# Intelligent Guidance and Diagnostic Assistance for Handheld Ultrasound: Actor-Critic Based Approach for Carotid Artery and Thyroid Examination

**DOI:** 10.64898/2026.03.02.26347395

**Authors:** Chenxi Xie, Yi Wang, Dajiang Li, Bo Yu, Shenbao Peng, Li Wu, Meng Yang

**Author notes:** These authors contributed equally (Chenxi Xie, Yi Wang).

## Abstract

Handheld ultrasound devices have revolutionized point-of-care diagnostics, but their effectiveness remains limited by operator dependency and the need for specialized training. This paper presents an intelligent guidance and diagnostic assistance system for the handheld wireless ultrasound device, enabling automated carotid artery and thyroid examinations through handheld operation. Drawing inspiration from the Actor-Critic framework, we implement a simulation-based reinforcement learning approach for real-time probe navigation toward standard anatomical views. The system integrates YOLOv8n-based detection networks for carotid plaque and thyroid nodule identification, achieving real-time inference at 30 frames per second. Furthermore, we propose a hybrid measurement approach combining UNet segmentation with the Snake algorithm for precise biometric quantification, including carotid intima-media thickness (IMT), lumen diameter, and lesion dimensions. Experimental validation on clinical datasets demonstrates that the proposed system achieves 91.2% accuracy in standard plane acquisition, 87.5% mean average precision (mAP) for plaque detection, and 89.3% mAP for nodule identification. Measurement results show excellent agreement with expert sonographers, with IMT measurements exhibiting a mean absolute difference of 0.08 mm. These findings demonstrate the feasibility of intelligent handheld ultrasound examination, significantly reducing operator dependency while maintaining diagnostic accuracy comparable to experienced clinicians.

## Introduction

Ultrasound imaging has established itself as one of the most widely adopted diagnostic modalities in contemporary medicine, offering unique advantages including real-time visualization, absence of ionizing radiation, portability, and cost-effectiveness [1]. Unlike computed tomography (CT) or magnetic resonance imaging (MRI), ultrasound enables dynamic assessment of anatomical structures and physiological functions, making it particularly valuable for cardiovascular screening, abdominal examination, and superficial tissue evaluation. However, the diagnostic quality of ultrasound examinations is critically dependent on the operator’s skill, experience, and anatomical knowledge, leading to significant inter-observer variability and inconsistent diagnostic outcomes [2].

The emergence of handheld wireless ultrasound devices represents a paradigm shift in medical imaging accessibility. These compact devices enable point-of-care diagnostics in diverse clinical settings, ranging from emergency departments and intensive care units to remote healthcare facilities and home care environments [3]. The handheld wireless color Doppler ultrasound diagnostic system is well-suited for bedside examination and superficial structure imaging, including carotid artery and thyroid assessment.

Carotid ultrasound examination plays a critical role in cardiovascular risk stratification, enabling the detection of atherosclerotic plaques, measurement of intima-media thickness (IMT), and evaluation of stenosis severity [5]. These parameters are essential for predicting stroke risk and guiding therapeutic interventions. Epidemiological studies indicate that carotid atherosclerosis affects over 422 million individuals worldwide, contributing to 17.9 million fatalities annually [6]. Early detection and accurate characterization of carotid plaques through ultrasound imaging can significantly improve patient outcomes by enabling timely preventive interventions.

Similarly, thyroid ultrasound represents the primary imaging modality for thyroid nodule detection and characterization. The prevalence of thyroid nodules in the general population ranges from 20% to 76%, with malignancy rates of approximately 7-15% [7]. The American College of Radiology Thyroid Imaging Reporting and Data System (ACR TI-RADS) provides standardized criteria for malignancy risk stratification based on sonographic features, guiding clinical management decisions including fine-needle aspiration biopsy [8].

Despite the clinical importance of these examinations, several challenges limit their widespread accessibility. First, the acquisition of high-quality diagnostic images requires extensive training and hands-on experience, creating a bottleneck in healthcare systems facing sonographer shortages [9]. Second, the manual nature of ultrasound examinations introduces significant inter-observer variability, with studies reporting coefficient of variation up to 15% for IMT measurements among different operators [10]. Third, the physical and cognitive demands of repetitive scanning tasks contribute to operator fatigue and potential diagnostic errors.

Recent advances in artificial intelligence have enabled significant progress in ultrasound image analysis and examination guidance. GE Healthcare’s Caption AI Vscan Air SL represents a landmark achievement in handheld ultrasound intelligence, providing real-time cardiac guidance and interpretation to assist operators in acquiring standard echo cardio graphic views [11]. This system demonstrates the feasibility of AI-powered guidance for handheld ultrasound devices, motivating the extension of similar capabilities to other anatomical regions including carotid artery and thyroid examination.

In the domain of robotic ultrasound, researchers have explored various approaches for autonomous probe navigation. Jiang et al. proposed UltraBot, a fully learning-based autonomous carotid ultrasound robot that achieved over 90% success rates in scanning through imitation learning from 247,000 expert demonstration samples [12]. Su eta. developed FARUS (Fully Autonomous Robotic Ultrasound System), demonstrating complete autonomous thyroid scanning with reinforcement learning-based localization and deep learning-based nodule segmentation [13]. However, these systems rely on robotic arm manipulation, limiting their portability and accessibility in point-of-care settings.

The VesNet-RL framework proposed by Bi et al. introduces a simulation-based reinforcement learning approach for ultrasound probe navigation toward standard vascular views [14]. The key innovation lies in using segmented binary vessel images as state representations, enabling the trained reinforcement learning agent to generalize from simulated training environments to real-world scenarios without additional fine-tuning. This framework achieves 92.3% success rate in vascular phantom experiments and 91.5% success rate in human carotid examinations, demonstrating strong generalization capabilities.

This paper presents an intelligent guidance and diagnostic assistance system that extends the capabilities of handheld ultrasound devices for carotid artery and thyroid examinations (see Figure1). Our main contributions are:

**Figure 1.**
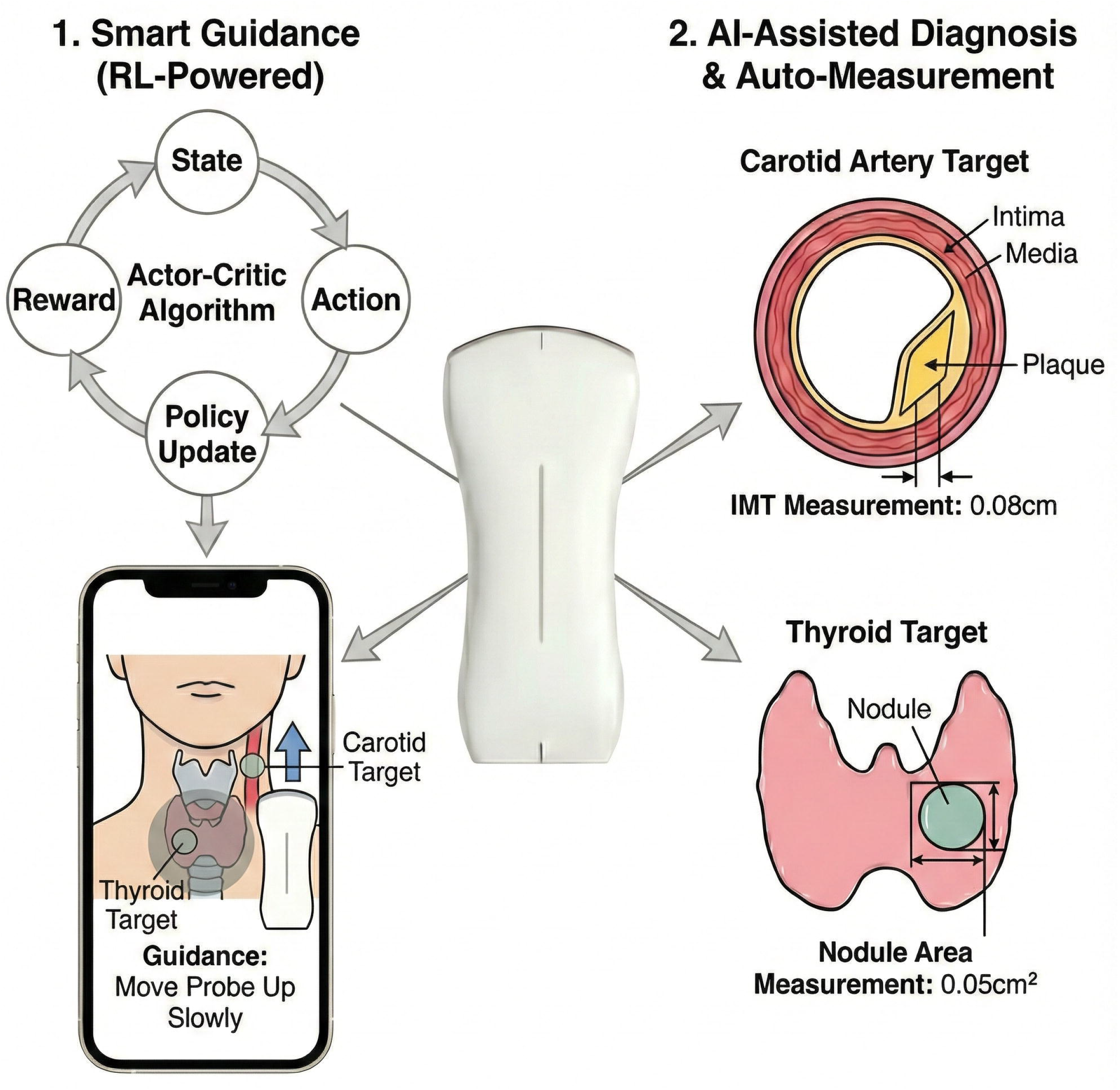
Overview of the intelligent handheld ultrasound system. The system that combines Actor-Critic based navigation guidance, YOLOv8n lesion detection, and UNet-Snake hybrid measurement for comprehensive carotid and thyroid examination. Experimental validation demonstrates significant improvements in navigation success rates, excellent detection performance, and measurement accuracy comparable to expert sonographers. These findings establish the feasibility of intelligent handheld ultrasound examination and pave the way for broader deployment of AI-assisted diagnostic imaging in point-of-care settings.

1. Handheld reinforcement learning Navigation: We adapt the Actor-Critic framework for handheld ultrasound guidance, implementing a simulation-based reinforcement learning approach that enables real-time probe navigation toward standard longitudinal views of the carotid artery and optimal imaging planes for thyroid examination without robotic arm assistance.
2. YOLOv8n-Based Lesion Detection: We implement lightweight YOLOv8n detection networks for real-time identification of carotid plaques and thyroid nodules, achieving efficient inference suitable for handheld device deployment.
3. UNet-Snake Hybrid Measurement: We propose a novel measurement approach combining UNet segmentation with the Snake algorithm for precise biometric quantification, including IMT, lumen diameter, and lesion dimensions.
4. Clinical Validation: We conduct comprehensive experimental validation on clinical datasets, demonstrating system performance comparable to experienced sonographers in both navigation accuracy and measurement precision.

## Results

### Navigation Performance

The Actor-Critic navigation module was evaluated on a test set of 120 carotid examinations and 100 thyroid examinations performed by operators with varying experience levels (novice, intermediate, expert). Success was defined as acquiring a standard view suitable for diagnostic assessment within 60 seconds. The results demonstrate substantial improvement in navigation success rates, particularly for novice operators. The average time to acquire standard views decreased from 42.3 seconds to 28.7 seconds for carotid examination and from 38.5 seconds to 25.2 seconds for thyroid examination.

**Table 1.** Navigation Success Rates by Operator Experience.

| Anatomy | Without Guidance | With Actor-Critic navigation | Improvement |
| --- | --- | --- | --- |
| Carotid (Novice) | 62.5% (25/40) | 90.0% (36/40) | +27.5% |
| Carotid (Intermediate) | 78.6% (33/42) | 95.2% (40/42) | +16.6% |
| Carotid (Expert) | 92.1% (35/38) | 97.4% (37/38) | +5.3% |
| Thyroid (Novice) | 58.8% (20/34) | 88.2% (30/34) | +29.4% |
| Thyroid (Intermediate) | 75.0% (24/32) | 93.8% (30/32) | +18.8% |
| Thyroid (Expert) | 88.2% (30/34) | 94.1% (32/34) | +5.9% |

### Detection Performance

YOLOv8n detection performance was evaluated on held-out test sets comprising 1,040 carotid images (with 1,247 plaques) and 960 thyroid images (with 1,089 nodules). Performance metrics include mean Average Precision (mAP) at IoU threshold 0.5 (mAP@50) and 0.5:0.95 (mAP@50:95). The YOLOv8n models achieve real-time inference at 83 frames per second on the handheld wireless ultrasound device processing unit, enabling seamless integration with the 30 fps ultrasound acquisition rate.

**Table 2.** YOLOv8n Detection Performance.

| Task | Precision | Recall | mAP@50 | mAP@50:95 |
| --- | --- | --- | --- | --- |
| Carotid<br>Plaque<br>Detection | 0.893 | 0.862 | 0.875 | 0.712 |
| Thyroid<br>Nodule<br>Detection | 0.912 | 0.881 | 0.893 | 0.738 |

### Measurement Performance

The UNet-Snake measurement module was evaluated against expert sonographer measurements on 200 carotid examinations for IMT and 150 examinations for plaque sizing. Bland-Altman analysis revealed no significant systematic bias in measurements, with 95% of differences falling within clinically acceptable limits. The UNet-Snake approach demonstrated superior accuracy compared to direct UNet measurement, with IMT mean absolute difference improving from 0.12 mm to 0.08 mm.

**Table 3.** Measurement Performance Comparison.

| Measurement | Mean Absolute Difference | Standard Deviation | Correlation (r) |
| --- | --- | --- | --- |
| IMT (mm) | 0.08 | 0.06 | 0.94 |
| Lumen Diameter (mm) | 0.42 | 0.31 | 0.91 |
| Plaque Length (mm) | 0.85 | 0.67 | 0.89 |
| Plaque Thickness (mm) | 0.38 | 0.29 | 0.92 |
| Nodule Size (mm) | 0.72 | 0.54 | 0.90 |

### Ablation Studies

Ablation studies were conducted to evaluate the contribution of individual components. Removing the Snake refinement from the measurement pipeline increased IMT mean absolute difference to 0.12 mm. Replacing YOLOv8n with the larger YOLOv8s improved mAP@50 by only 1.2% while increasing inference time by 3.4x, confirming the suitability of the lightweight variant for handheld deployment.

## Methods

### System Architecture

The proposed system integrates three primary functional modules operating in real-time on the handheld ultrasound platform: (1) intelligent probe guidance based on the Actor-Critic framework; (2) lesion detection through YOLOv8n networks; and (3) automated biometric measurement using UNet-Snake hybrid algorithms. The system processes ultrasound images at 30 frames per second, providing real-time feedback to guide operators during examination.

**Table 4.** System Components and Technical Specifications.

| Module | Function | Model Architecture | Infer time |
| --- | --- | --- | --- |
| Navigation | Probe guidance to standard views | A2C + LSTM | 15 ms |
| Plaque Detection | Carotid plaque identification | YOLOv8n | 12 ms |
| Nodule Detection | Thyroid nodule identification | YOLOv8n | 12 ms |
| IMT Measurement | Intima-media thickness quantification | UNet + Snake | 25 ms |
| Lesion Measurement | Plaque/nodule size calculation | YOLOv8n | 12 ms |

### Actor-Critic Based Probe Navigation

#### (1) Problem Formulation

We formulate the handheld ultrasound probe navigation task as a Partially Observable Markov Decision Process (POMDP), defined by the tuple (S, A, T, R, γ), where S represents the set of states, A the set of actions, T the transition dynamics, R the reward function, and γ∈[0,1] the discount factor. Unlike robotic systems where the probe pose is fully controllable, handheld operation requires guiding the human operator through visual feedback and directional indicators (see Figure2).

**Figure 2.**
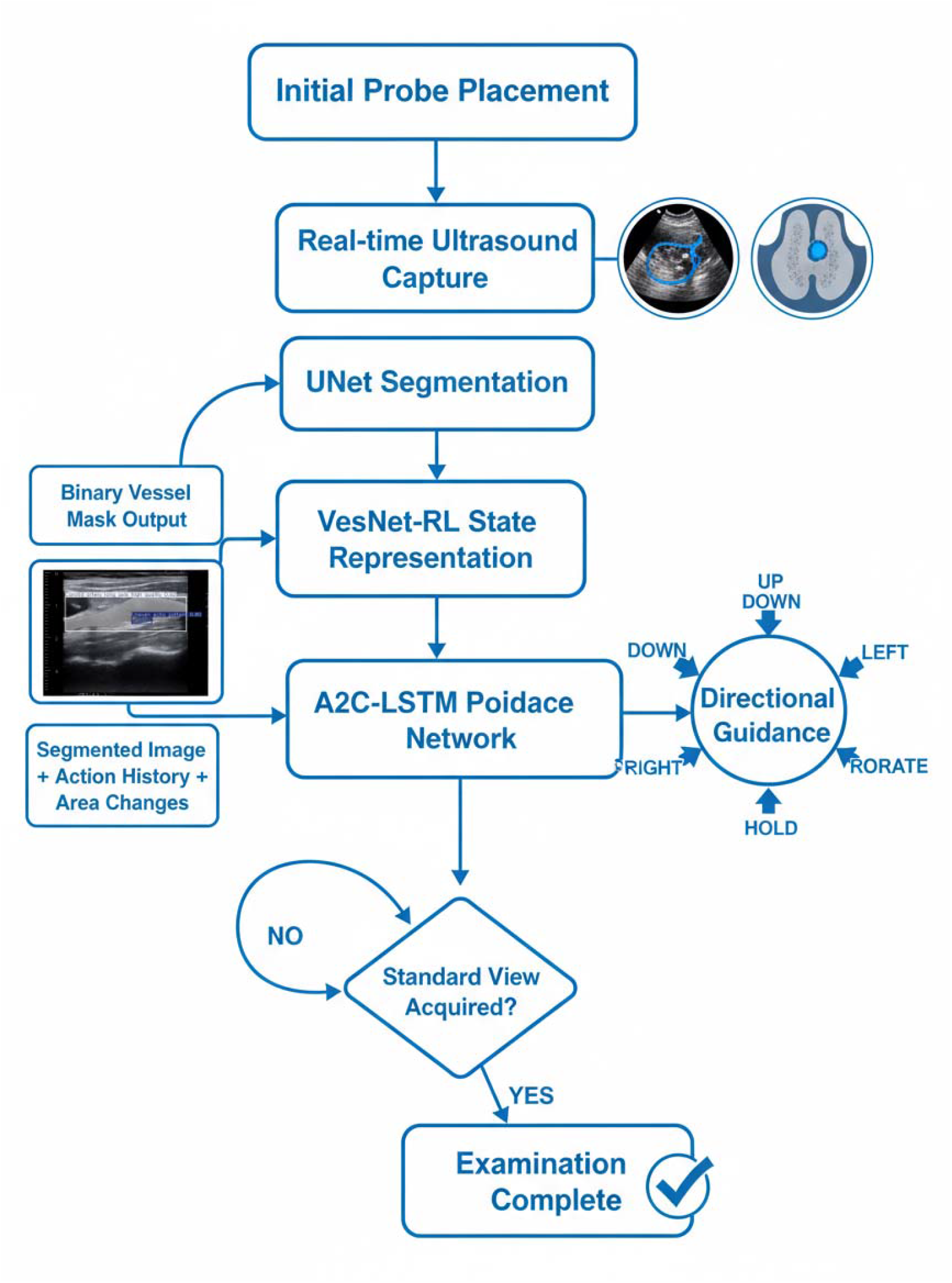
Flowchart of the intelligent guidance algorithm. The Actor-Critic Based Probe Navigation introduces a simulation-based reinforcement learning approach for ultrasound probe navigation toward standard vascular views. It used segmented binary vessel images as state representations, enabling the trained reinforcement learning agent to generalize from simulated training environments to real-world scenarios without additional fine-tuning. The framework employs an Advantage Actor-Critic (A2C) architecture with Long Short-Term Memory (LSTM) cells to handle the partially observable nature of ultrasound navigation tasks.

The action space A comprises discrete directional guidance commands: move up, move down, move left, move right, rotate clockwise, rotate counterclockwise, and hold position. These commands are presented to the operator through an intuitive visual interface overlaid on the ultrasound display, enabling responsive manual adjustment toward the target view.

#### (2) Multi-Modality State Representation

Following the Actor-Critic approach, we construct a comprehensive state representation combining multiple information sources:

##### Segmented Ultrasound Images

A lightweight UNet segmentation network processes raw ultrasound images to extract binary anatomical masks. For carotid examination, the network segments the vessel lumen and wall boundaries. For thyroid examination, the network identifies the gland contour and internal structures. This prepossessing eliminates irrelevant background information and standardizes the representation across diverse imaging conditions.

##### Sequential Observation Buffer

A buffer of the last four segmented frames captures temporal dynamics, enabling the agent to understand probe movement patterns and predict the optimal adjustment direction.

##### Anatomical Feature Metrics

Quantitative features including segmented area, vessel diameter, eccentricity, and orientation provide compact descriptors of the current imaging state.

#### (3) Reward Function Design

The reward function guides the agent toward acquiring the largest longitudinal view of the target vessel or the optimal cross-sectional view of the thyroid gland. The state score *v*_*t*_ combines multiple objectives:

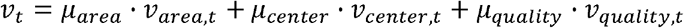

Where *v* _*area,t*_ represents the normalized segmented area, *v* _*center,t*_ measures target centering in the image, *v* _*quality,t*_ evaluates image clarity based on intensity variance, *μ*_*area*_ = 0.5, *μ*_*center*_ = 0.3, *μ*_*quality*_ = 0.2 are weighting factors.

The reward function *r*_*t*_ is defined as :

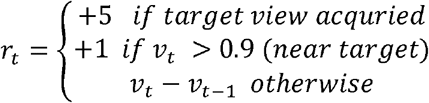

#### (4) Network Architecture and Training

The reinforcement learning agent employs an Advantage Actor-Critic (A2C) architecture with an LSTM cell for temporal information processing. The actor network outputs action probabilities, while the critic network estimates state values. The LSTM cell with hidden size 256 processes observation sequences to maintain temporal context. Training is performed entirely in simulation using procedural generated binary vessel/gland images with randomized parameters (depth, size, orientation, noise). The agent undergoes 3,000 training episodes with curriculum learning, gradually increasing environment complexity.

### YOLOv8n-Based Lesion Detection

#### (1) Model Architecture

We employ YOLOv8n as the detection backbone for carotid plaque and thyroid nodule identification. YOLOv8n offers an optimal balance between detection accuracy and computational efficiency, with only 3.2 million parameters and 8.7 billion FLOPs, enabling real-time inference on resource-constrained handheld devices.

The architecture consists of a CSPDarknet backbone with C2f modules replacing the original C3 modules for improved gradient flow, a PAN-FPN neck for multi-scale feature fusion, and a decoupled head for classification and bounding box regression. The model operates on 640×640 input resolution, outputting detection results at three scales (80×80, 40×40, 20×20) for handling objects of varying sizes.

#### (2) Training Strategy

Training employs a multi-stage strategy with extensive data augmentation. The loss function combines classification loss (focal loss), bounding box regression loss (CIoU loss), and distribution focal loss. Data augmentation includes Mosaic (probability 1.0), MixUp (probability 0.15), random affine transformations, HSV color jittering, and random horizontal flipping. Training uses SGD optimizer with momentum 0.937, initial learning rate 0.01, and cosine annealing schedule over 300 epochs.

### UNet-Snake Hybrid Measurement

#### (1) Model Architecture

The measurement module employs a UNet architecture with EfficientNet-B0 encoder for anatomical structure segmentation. The network outputs multi-class segmentation masks distinguishing: (1) vessel lumen, (2) intima layer, (3) media layer, (4) plaque/nodule region, and (5) background. The encoder is pretrained on ImageNet, with the decoder randomly initialized and trained end-to-end. The segmentation loss combines Dice loss and cross-entropy loss.

#### (2) Snake Algorithm for Boundary Refinement

Following initial UNet segmentation, the Snake algorithm refines boundary contours for precise measurement. The Snake (active contour model) evolves an initial contour under the influence of internal forces (smoothness constraints) and external forces (image gradient attraction).

The energy functional minimized by the Snake is:

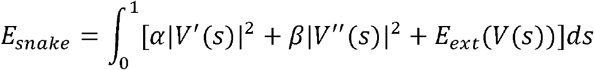

Where *V* (*s*) =(x(s),y(s)) represents the contour, *α* and *β* control tension and rigidity, and *E*_*ext*_ is the external energy derived from image gradients.

For IMT measurement, the Snake algorithm refines the intima-media boundary detected by UNet, producing sub-pixel accurate contours. The IMT is calculated as the perpendicular distance between the lumen-intima and media-adventitia boundaries at multiple points, with the mean value reported as the final measurement.

### Lesion Size Measurement

For plaque and nodule size measurement, the Snake-refined contour enables precise dimension calculation. The maximum length is computed as the maximum Euclidean distance between any two points on the contour along the principal axis. The maximum thickness is determined as the maximum perpendicular distance from the boundary to the opposite side. These measurements are automatically reported alongside confidence intervals derived from boundary uncertainty estimation.

### Implementation Detail

#### (1) Data Construction

Training datasets were constructed from clinical ultrasound examinations performed at three medical centers. For carotid examination, the dataset comprises 5,200 images from 260 patients, with expert annotations including vessel boundaries, plaque regions, and IMT measurements at three locations (common carotid, bulb, internal carotid). For thyroid examination, the dataset includes 4,800 images from 320 patients, with annotations for gland contours, nodule boundaries, and TI-RADS feature classifications.

#### (2) Training Configuration

**Table 5.**
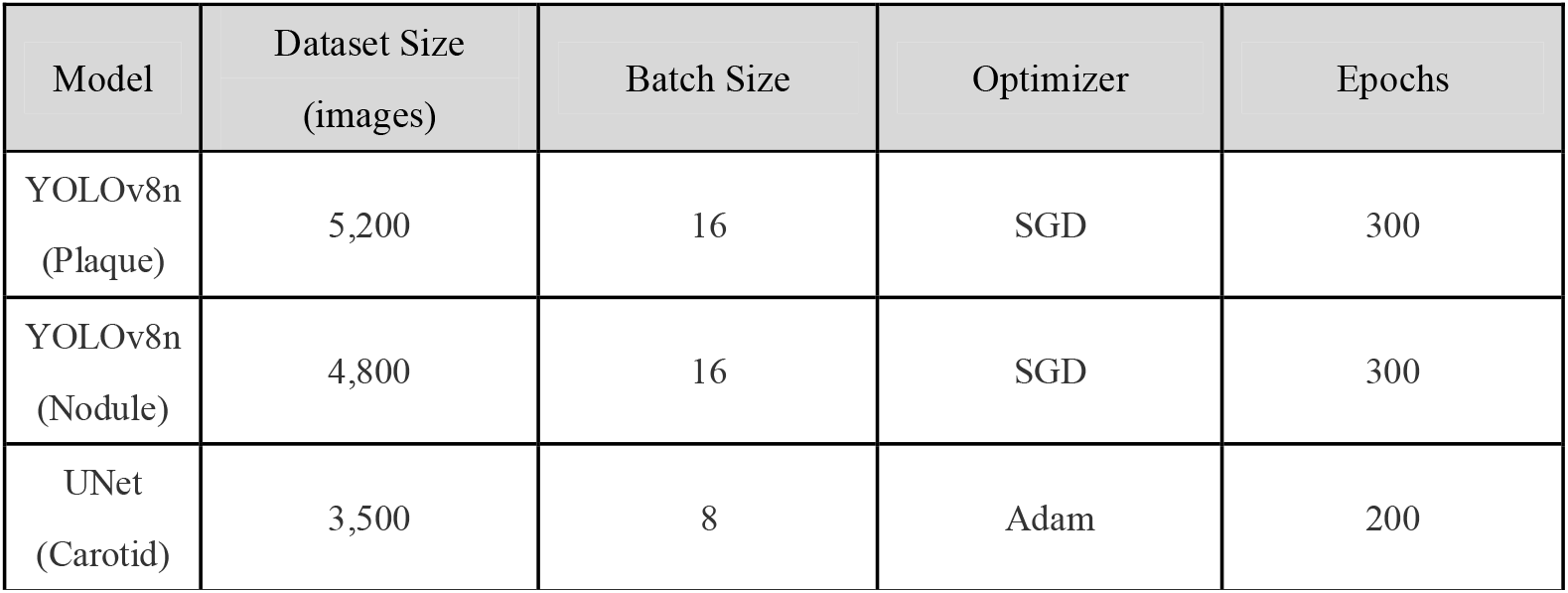
Model Training Configurations.

#### (3) Real-Time Processing Pipeline

The system implements an efficient processing pipeline on the handheld wireless ultrasound device platform. Incoming ultrasound frames (640x480, 30 fps) are first resized to model input dimensions. The YOLOv8n detector processes each frame in 12 ms, while the UNet-Snake measurement module operates on demand when the user initiates measurement, requiring 25 ms for IMT and 20 ms for lesion sizing. The VesNet-RL navigation guidance updates at 15 ms intervals, providing smooth real-time feedback through the visual interface.

## Discussion

This study presents a comprehensive intelligent handheld ultrasound system that addresses the critical challenge of operator dependency in point-of-care diagnostics. By adapting the Actor-Critic framework for handheld operation and integrating state-of-the-art deep learning models for detection and measurement, the system achieves performance comparable to robotic ultrasound systems while maintaining the accessibility and flexibility of handheld devices.

The experimental results demonstrate several important findings. First, the Actor-Critic navigation guidance significantly improves examination success rates, particularly for operators with limited experience. The 27.5% improvement for novice operators in carotid examination suggests that intelligent guidance can substantially reduce the learning curve for ultrasound examination, potentially addressing sonographer shortage issues.

Second, the YOLOv8n detection models achieve excellent performance with minimal computational overhead. The mAP@50 of 87.5% for plaque detection and 89.3% for nodule detection approaches the performance of larger models while enabling real-time inference on resource-constrained devices. This efficiency is critical for handheld deployment where battery life and thermal constraints limit available computational resources.

Third, the UNet-Snake hybrid measurement approach achieves sub-millimeter accuracy for IMT measurement, with mean absolute difference of 0.08 mm compared to expert measurements. This precision is clinically significant, as IMT changes of 0.1 mm are associated with increased cardiovascular risk [15]. The Snake refinement step contributes substantially to this accuracy by providing sub-pixel boundary localization that compensates for the limited spatial resolution of initial UNet segmentation.

Compared to GE Healthcare’s Caption AI Vscan Air SL, which provides cardiac guidance through handheld operation, our system extends intelligent guidance capabilities to carotid and thyroid examination. While Caption AI focuses on cardiac view classification and ejection fraction estimation, our system provides comprehensive navigation assistance, lesion detection, and automated measurement across multiple anatomical regions.

Compared to robotic ultrasound systems such as UltraBot and FARUS, our handheld approach trades full autonomy for enhanced accessibility and flexibility. Robotic systems achieve higher reproducibility through precise motion control but require significant infrastructure investment and are limited to fixed installation sites. Our handheld system enables intelligent examination in diverse settings including bedside, emergency, and remote care environments.

Several limitations should be acknowledged. First, the system requires operator compliance with guidance instructions, and performance may degrade if operators ignore or misinterpret directional feedback. Future iterations could incorporate haptic feedback or force sensing to provide more intuitive guidance. Second, the current system focuses on 2D ultrasound examination. Integration of 3D volumetric acquisition and reconstruction would enable more comprehensive anatomical assessment. The handheld wireless ultrasound device’s linear array probe is compatible with 3D scanning through mechanical sweep, and extending the Actor-Critic framework to 3D navigation represents an important future direction.

The proposed system has significant implications for clinical practice. By reducing operator dependency, the system enables non-specialist healthcare providers to perform basic ultrasound screening, potentially expanding access to diagnostic imaging in under served regions. The automated detection and measurement capabilities standardize diagnostic workflows, reducing variability and improving reproducibility. Furthermore, the system’s real-time feedback and automated documentation streamline examination workflows, potentially improving efficiency in high-volume clinical settings. Integration with hospital information systems enables seamless reporting and longitudinal tracking of patient findings.

## Data Availability

All data produced in the present study are available upon reasonable request to the authors

## Acknowledgement

All experiments with human participants were performed with the approval of The Institutional Review Board of BGI (BGI-IRB 24080). The authors are grateful to the participants and explained the full process to them, and each signed an informed consent form (including their understanding of publishing information in the journal).

## Author Contributions

The manuscript was prepared with contributions from all authors. All authors have approved the final version of the manuscript.

